# Elevated lysophosphatidylcholines during SSRI-induced neural differentiation correlate with early neurodevelopmental symptoms

**DOI:** 10.1101/2025.05.30.25328657

**Authors:** Abishek Arora, Kristina Vacy, Cátia Marques, Mihai-Ovidiu Degeratu, Francesca Mastropasqua, Jenny Humphrey, Xuan Ye, Marika Oksanen, the Barwon Infant Study Investigator Group, Peter Vuillermin, Anne-Louise Ponsonby, Ingela Lanekoff, Kristiina Tammimies

**Affiliations:** Center of Neurodevelopmental Disorders (KIND), Centre for Psychiatry Research, Department of Women’s and Children’s Health, Karolinska Institutet, and Child and Adolescent Psychiatry, Stockholm Health Care Services, Region Stockholm, Stockholm, Sweden; Astrid Lindgren Children’s Hospital, Karolinska University Hospital, Region Stockholm, Stockholm, Sweden; SciLifeLab, Department of Women’s and Children’s Health, Karolinska Institutet, Stockholm, Sweden; Florey Institute of Neuroscience and Mental Health, University of Melbourne, Victoria, Australia; Melbourne School of Population and Global Health, University of Melbourne, Victoria, Australia; Department of Chemistry - BMC, Uppsala University, Uppsala, Sweden; Murdoch Children’s Research Institute, Royal Children’s Hospital, and Department of Paediatrics, University of Melbourne, Victoria, Australia; Center of Excellence for the Chemical Mechanisms of Life, Uppsala University, Uppsala, Sweden

**Keywords:** Selective serotonin reuptake inhibitors, induced pluripotent stem cells, clinical cohort, neurodevelopmental disorders, metabolomics, cellular assays

## Abstract

Selective serotonin reuptake inhibitors (SSRIs) are often prescribed during pregnancy. Yet, epidemiological studies link *in-utero* SSRI exposure with neurodevelopmental disorders, such as autism and ADHD. The potential molecular mechanisms by which SSRIs impact early neurodevelopment are not fully understood. We exposed neuroepithelial stem cells derived from four human induced pluripotent stem cells lines to fluoxetine, citalopram, sertraline, and paroxetine. We then assessed cellular viability, reactive oxygen species (ROS) levels, mitochondrial function using adenosine triphosphate (ATP) assays, and performed high-throughput metabolomics at two timepoints: proliferation and neural differentiation stages. The key metabolic findings were validated in the *in-vitro* model and in a complementary population-based cohort, the Barwon Infant Study, consisting of 1074 mother-child pairs with analysed cord-blood metabolomes. Sertraline and paroxetine significantly decreased ROS and ATP levels, indicating mitochondrial alteration. Metabolomic profiling revealed consistent elevation of three lysophosphatidylcholines (LPCs 16:0, 18:0, 18:1) across all SSRIs except citalopram. We further observed elevated LPC levels in the cord blood of infants prenatally exposed to SSRIs, with a dose-dependent correlation to autism and ADHD-related symptoms at age two. Furthermore, the three LPCs modulated ROS and ATP levels in the neural cells. These findings provide insights into SSRI-induced molecular changes, highlight candidate metabolites that may warrant further investigation as indicators of SSRI exposure, and emphasise the need for exploring prenatal SSRI exposure effects and neurodevelopmental outcomes.

## INTRODUCTION

Selective serotonin reuptake inhibitors (SSRIs) are widely prescribed antidepressants that block the reuptake of serotonin by binding to the serotonin transporter, resulting in elevated levels of serotonin in the synaptic cleft.^1^ Depression affects ∼28% of pregnant women^2^ and SSRI use during pregnancy has increased over recent decades.^3^ Therefore, it is critical to understand their impact on foetal development. Although considered relatively safe, several epidemiological studies suggest that SSRI exposure during pregnancy may increase the likelihood of neurodevelopmental disorders (NDDs), including autism and attention-deficit/hyperactivity disorder (ADHD). For instance, a meta-analysis of 18 studies showed that *in-utero* exposure to SSRIs increases the likelihood of autism (hazard ratio, HR=1.27; 95% confidence interval, CI=1.10–1.47).^4^ Nevertheless, several confounding factors, including shared genetic liability factors, could still influence this.^5,6^ Beyond NDDs, a recent review has found a strong association of different SSRIs with several adverse foetal health outcomes,^7^ including an increased burden of cardiovascular complications in neonates.^8,9^

Sertraline, citalopram, paroxetine, and fluoxetine are the most commonly prescribed SSRIs to women of reproductive age.^10,11^ These SSRIs vary in pharmacological properties such as half-life (approximately 21 hours to a few days), the relationship between dose/blood levels and the dose range of prescription.^12^ SSRIs can cross the placenta^13^ and subsequently alter foetal serum serotonin levels,^14^ with a plausible impact on the developing brain.^15^ Neurodevelopment at the early stages of gestation, prior to the formation of the intact blood-brain barrier^16^ and widespread synaptic connectivity, relies on regulation by neurotransmitters.^17^ Serotonergic activity is one of the building blocks of neuronal organisation, with serotonin being one of the first neurotransmitters to emerge.^18^ Although many uncertainties remain around SSRIs and their mechanism of action, few studies have demonstrated wide-ranging cellular effects, including an alteration in reactive oxygen species (ROS) levels^19^ and an impact on mitochondrial oxidative phosphorylation.^20^ During neurodevelopment, ROS functions as a signalling molecule contributing to neuronal growth and synaptogenesis.^21^ However, excessive ROS-induced oxidative stress can lead to genetic mutations, lipid peroxidation, and mitochondrial dysfunction.^22^ Furthermore, in a previous study, we demonstrated that fluoxetine exposure alters lipid and energy metabolism, as observed in the transcriptome and metabolome of differentiating neuroepithelial stem (NES) cells.^23^ These findings, together with evidence that SSRIs accumulate in cellular membranes and can perturb phospholipid composition,^24^ give clear indications that there are cellular metabolic effects of SSRIs. Despite these efforts, there are no systematic studies investigating the potential molecular mechanisms of SSRI exposure in early neuronal development and how these could link to NDDs.

In this study, we hypothesised that SSRI exposure could alter markers of oxidative stress and energy metabolism in developing neural cells, lead to consistent lipid signatures across multiple SSRIs, and that these signatures would also associate with SSRI exposure and early neurodevelopmental traits in a clinical birth cohort.

To test these hypotheses, we used human induced pluripotent stem cell (iPSC)- derived NES cells undergoing undirected differentiation into neurons and exposed them to four commonly prescribed SSRIs. We, first, evaluated the effects of SSRI exposure on cellular viability, ROS, and oxidative phosphorylation of ATP. After characterising these *in-vitro* effects, we performed untargeted mass spectrometry analysis to develop metabolomic profiles of the general and specific effects of each SSRI.

Finally, we analysed the top three upregulated lipids from the cellular models in cord blood from a population-based cohort, the Barwon Infant Study (BIS), consisting of 1074 mother-child dyads, and investigated the association between SSRI exposure, the cord blood lipid levels and correlation with autism and ADHD traits. This integrated approach aims to advance understanding of SSRI-mediated effects during pregnancy, including their potential impact on neural cell development, differentiation, and associated metabolic pathways, and to inform future epidemiological and clinical evaluations.

## METHODS

### Cell culture

For the purpose of this study, previously generated and described human iPSC lines from tissue samples of a male: CTRL-9-II – CTRL_Male_^25^ and female: AF22 – CTRL_Female_^26^ neurotypical donors and two male donors with known genetic variants for NDDs including autism: HNRNPU_del/+•_ – ASD_HNRNPU_^27^ and ASD_CASK_SS_ – ASD_CASK,_^28^ were used (Supplementary Table S1). Information pertaining to the generation and quality control of the iPSC lines, as well as their relevant clinical background are provided in the cited publications. These lines were selected to capture conserved SSRI exposure effects across sex and neurodevelopmental genetic backgrounds rather than to model population-level heterogeneity or genotype-specific responses. The iPS cells were differentiated into neural progenitor state and maintained as NES cells, as previously described.^26,29^ While NES cells are neural progenitors with both neurogenic and gliogenic potential, upon differentiation they are primarily neurogenic unless they are specifically differentiated to glia.^26,29^ The iPSC lines and this NES-to-neuron differentiation system have been previously validated and characterised for pluripotency, neural progenitor identity and neuronal differentiation in multiple independent studies and were therefore not re-characterised in the present work.^25–28^

The NES cells were seeded (40,000 cells/cm^2^) on tissue culture treated plates (Sarstedt), coated with 20 μg/mL poly-L-ornithine (P3655, Sigma-Aldrich) and 1 μg/mL Laminin2020 (L2020, Sigma-Aldrich). These were grown in DMEM/F-12 Glutamax basal medium (31331-093, Thermo Scientific) supplemented with 0.05x B27 (17504044, Thermo Scientific), 1x N-2 (17502001, Thermo Scientific), 10 U/ml Penicillin-Streptomycin (15140122, Thermo Scientific), 10 ng/mL recombinant human bFGF (13256-029, Thermo Scientific) and 10 ng/mL recombinant human EGF (AF-100-15, Peprotech). The undirected differentiation of NES cells to neural cell population was achieved by the withdrawal of growth factors from the culture medium.^26,29^ Here, the NES cells were again seeded (40,000 cells/cm^2^) on tissue culture treated plates (Sarstedt), coated with 20 μg/mL poly-L-ornithine (P3655, Sigma-Aldrich) and 1 μg/mL Laminin2020 (L2020, Sigma-Aldrich). However, these were grown in DMEM/F-12 Glutamax basal medium (31331-093, Thermo Scientific) supplemented with 0.5x B27 (17504044, Thermo Scientific), 1x N-2 (17502001, Thermo Scientific) and 10 U/ml Penicillin-Streptomycin (15140122, Thermo Scientific). The cells were differentiated for 5 days or 28 days post induction of neural differentiation, followed by sampling based on the downstream application. Day 5 represents an early neuronal progenitor differentiation stage, whereas day 28 reflects a more mature neuronal lineage enriched for post-mitotic neurons in this NES-based differentiation system.^25–28^

### Selective serotonin reuptake inhibitors (SSRIs)

We evaluated the effects of fluoxetine hydrochloride (FH; F132, Sigma-Aldrich), citalopram hydrobromide (CH; C7861, Sigma-Aldrich), sertraline hydrochloride (SH; S6319, Sigma-Aldrich) and paroxetine hydrochloride (PH; P9623, Sigma Aldrich) in our cell lines. SSRI stock solutions of 0.01 M concentration were prepared in absolute ethanol (SH) or MilliQ H_2_O (FH, CH and PH). Working solutions were prepared in MilliQ H_2_O at 0.001 M. For the *in-vitro* exposures at the required concentration(s), the SSRI working solutions were subsequently mixed with DMEM/F-12 Glutamax basal medium (31331-093, Thermo Scientific) supplemented with 0.5x B27 (17504044, Thermo Scientific), 1x N-2 (17502001, Thermo Scientific) and 10 U/ml Penicillin-Streptomycin (15140122, Thermo Scientific). Vehicle controls were also prepared, as required. Freshly prepared exposure medium was replaced on every second day during the period of neural differentiation.

### Cytotoxicity assay

To select the appropriate treatment concentration of the SSRIs for our cell lines, we performed cytotoxicity testing. The treatment concentration ranges (0.00–30.00 µM) for the SSRIs were determined based on those optimised in our previous study using FH,^23^ followed by testing in the CTRL_Male_ cell line using the MTS assay (G3580, Promega).

The NES cells were seeded (12,000 cells/well) in 96-well plates (Sarstedt) and exposed to a concentration range of the SSRI being evaluated. Growth factors were withdrawn from the cell culture, to initiate neural differentiation. The exposure media was changed every second day. At 24- and 120-hours (5 days) post exposure, the cells were treated with MTS (3-(4,5-dimethylthiazol-2-yl)-5-(3-carboxymethoxyphenyl)-2-(4-sulfophenyl)-2H-tetrazolium) reagent, based on instructions from the manufacturer. After 3 hours of incubation at 37°C and 5% CO_2_, absorbance was recorded at 490 nm using a spectrophotometric microplate reader (FLUOstar Omega, BMG Labtech). For this assay, one independent experiment was performed at each timepoint, with a suitable number of technical replicates as visualised in the plots.

Quantification of percentage viability at each concentration and timepoint was done in R (v4.1.2).^30^ The response was represented as a regression line generated using the Loess method for local regression fitting, and the shaded area denoted the 95% confidence interval. Two-way ANOVA, followed by Tukey’s post-hoc correction for multiple comparisons, was used to test for significance (adj. p<0.05) of the reported findings.

### Cellular ROS assay

The cellular ROS assay (ab113851, Abcam) was used to determine total ROS levels at day 5 and 28 of SSRI exposures (FH, CH and PH: 3.0 µM; SH: 1.5 µM). In short, NES cells were seeded (12,000 cells/well) in dark walled, clear bottom 96-well plates (Sarstedt) and allowed to adhere overnight at 37°C, 5% CO_2_ incubation. Thereafter, neural differentiation was induced by withdrawing growth factors from the culture medium accompanied by exposure to the SSRIs, with the culture medium being changed on every alternative day.

The culture medium was aspirated, and the cells were exposed to the fluorogenic dye, DCFDA (2’,7’-dichlorodihydrofluorescein diacetate), at a concentration of 10 μM, in 100 μL of 1X Buffer (from the kit) and incubated at 37°C, 5% CO_2_ for 45 minutes. Fluorescence at excitation wavelength 485 nm and emission wavelength 520 nm was recorded using a spectrophotometric microplate reader (FLUOstar Omega, BMG Labtech). For these assays, two independent experiments were performed for each differentiation timepoint, with a suitable number of technical replicates as visualised in the plots. All readings were reported as percentage of control. Normality testing was performed using the Shapiro-Wilk Test. Subsequently, Kruskal-Wallis rank sum test, followed by Nemenyi’s post-hoc correction for multiple comparisons, was used to test for the significance (adj. p<0.05) of the reported findings in R (v4.1.2).^30^

### Mitochondrial ToxGlo assay

To test and correct for the Crabtree effect,^31^ the mitochondrial ToxGlo assay (G8000, Promega) was used to check for mitochondrial vulnerability, with effects on oxidative phosphorylation and subsequent ATP generation, to the potent mitotoxin sodium azide.^32^ In short, NES cells were seeded (12,000 cells/well) in white walled, clear bottom 96-well plates (Greiner) and allowed to adhere overnight at 37°C, 5% CO_2_ incubation. The assay was used to determine cell membrane integrity and adenosine triphosphate (ATP) levels after 24 hours, in accordance with the optimised manufacturer’s instructions (Promega).

On the next day, the culture medium was replaced with glucose free DMEM (11966025, Thermo Scientific) supplemented with 10 mM galactose (G5388, Sigma-Aldrich), instead of DMEM-F12 Glutamax basal medium (31331-093, Thermo Scientific), 90 minutes prior to the assay. The galactose induced the cells to primarily rely on oxidative phosphorylation to produce ATP, instead of glycolysis. Cell membrane integrity was assessed by measuring a distinct protease activity associated with necrosis using a fluorogenic peptide substrate, bis-AAF-R110. After incubation at 37°C, 5% CO_2_ for 30 minutes, fluorescence at excitation wavelength 485 nm and emission wavelength 520 nm was recorded using a spectrophotometric microplate reader (FLUOstar Omega, BMG Labtech).

Next, the ATP detection reagent was added, causing cell lysis, and producing a luminescent signal proportional to the amount of total ATP present. Immediately after, luminescence was recorded using a spectrophotometric microplate reader (FLUOstar Omega, BMG Labtech). For this assay, one independent experiment was performed, with a suitable number of technical replicates as visualised in the plots. All readings were reported as percentage of control. Two-way ANOVA followed by Tukey’s post-hoc correction for multiple comparisons, was used to test for significance (adj. p<0.05) of the reported findings in R (v4.1.2).^30^

### Cellular ATP assay

The CellTiter Glow 2.0 assay (G9242, Promega) was used to quantify the total ATP levels generated by oxidative phosphorylation in response to the SSRI exposures. This was done at day 5 (FH, CH and PH: 3.0 µM; SH: 1.5 µM) and day 28 of SSRI exposures (FH, CH, SH and PH: 1.5 µM). In short, NES cells were seeded (12,000 cells/well) in white walled, clear bottom 96-well plates (Greiner) and allowed to adhere overnight at 37°C, 5% CO_2_ incubation. Thereafter, neural differentiation was induced by withdrawing growth factors from the culture medium and exposure to SSRIs, with the culture medium being changed at every alternative day.

On day 5 and day 28 post induction of neural differentiation, the culture medium (100 µL/well) was replaced with glucose free DMEM (11966025, Thermo Scientific) supplemented with 10 mM galactose (G5388, Sigma-Aldrich), instead of DMEM-F12 Glutamax basal medium (31331-093, Thermo Scientific), 90 minutes prior to the assay. Thereafter, the cells were incubated at room temperature for 30 minutes followed by the addition of the CellTiter Glo 2.0 reagent (100µL/well) and mixing on an orbital shaker for 2 minutes to induce cell lysis. The cells were then incubated at room temperature for another 10 minutes followed by recording luminescence using a spectrophotometric microplate reader (FLUOstar Omega, BMG Labtech). For these assays, two independent experiments were performed for each differentiation timepoint, with a suitable number of technical replicates as visualised in the plots.

All readings were reported as percentage of control. For the day 5 results, normality testing was performed using the Shapiro-Wilk Test. Subsequently, Two-way ANOVA followed by Tukey’s post-hoc correction for multiple comparisons, was used to test for significance (adj. p<0.05) of the reported findings. For the day 28 results, normality testing was also performed using the Shapiro-Wilk Test. Subsequently, the Dunn (1964) Kruskal-Wallis multiple comparison with p-values adjusted with the Holm method was used to test for significance (adj. p<0.05) of the reported findings in R (v4.1.2).^30^

### Direct infusion electrospray ionisation mass spectrometry (ESI-MS)

Untargeted metabolomic profiling was performed using the direct infusion probe (DIP) for ESI-MS^33^ on snap-frozen cells at day 5 (N=120) and day 28 (N=120) of SSRI exposures during neural differentiation of NES cells, with two biological replicates having three technical replicates each for the treated and untreated groups in all four cell lines: CTRL_Male_, CTRL_Female_, ASD_HNRNPU_ and ASD_CASK_. For the day 5 samples, the cells were exposed to 3.0 µM of FH, CH and PH, and 1.5 µM of SH, as per the previously determined concentrations based on the cytotoxicity assays (Supplementary Fig. S1A). For the day 28 samples, the cells were exposed to a standardised dose of 1.5 µM to enable direct comparison of long-term exposure related metabolomic effects induced by each SSRI.

The cell culture plates were stored at -80°C prior to lysing with the electrospray solvent containing 9:1 methanol:water, with 0.1% formic acid and 28 internal standards. To enable comparisons, all cell samples were normalized to total cell counts per sample. The samples were analysed on a QExactive^TM^ Orbitrap instrument (Thermo Scientific) at mass resolving power of 140,000 (*m/Δm*) using untargeted full scanning between 70 and 1000 Da in positive mode. The resulting data was extracted using 30 seconds data per sample and sorted in MZmine 2.^34^ One-point quantification of metabolite concentrations in the sample was calculated using the intensity ratio of endogenous metabolite to the internal standard and multiplied with the concentration of the internal standard.^33^

### Differential metabolomics and feature selection

Principal component analysis (PCA) was performed in R (v4.1.2)^30^ followed by visualisation with the *factoextra* package (v1.0.7) (https://cran.r-project.org/package=factoextra). Prior to the PCA, any missing values (0.03% of the dataset) were imputed using the *missMDA* package (v1.18)^35^ in R (v4.1.2).^30^ Here, the number of dimensions of the PCA were estimated by cross-validation using the k-fold method followed by imputation of missing values using the PCA model.

To test for significance of the resulting metabolite concentrations (µM) following the mass spectrometry analysis workflow, a mixed linear model using the *nlme* package (v3.1-162)^36^ in R (v4.1.2)^30^ was applied to the dataset for each SSRI (FH, CH, SH and PH) at each timepoint (day 5, day 28). Any missing values were treated as *NA* values. Here, the mixed linear model used was *lme(concentration ∼ exposure, random = ∼ 1 | cell line)*. The data was filtered prior to the application of the mixed linear model, based on the SSRI exposure. The obtained p values were adjusted for multiple comparisons using the FDR method in R (v4.1.2).^30^ Significantly modulated metabolites (Treatment vs. Control) were selected based on a significance threshold of adjusted p<0.05 and were visualised using the *ggplot2* package (v3.3.5)^37^ in R (v4.1.2).^30^

Feature selection of shared SSRI effects on the metabolites was performed using the statistical meta-analysis module of MetaboAnalyst 5.0^38^. Metabolite concentration lists were provided for each SSRI at day 5 and day 28, respectively. Followed by performing an automated data integrity check, normalisation was performed using log2 transformation and data-scaling. Data scaling adjusts each variable/feature by a scaling factor computed based on the dispersion of the variable, in this case auto-scaling was used (mean-centred and divided by the standard deviation of each variable). For the selection of significant features that were shared by all exposure conditions, p values from each analysis were combined using the Fisher’s method, -2*∑Log(p). A combined *T statistic* and combined p value was provided for each metabolite (feature), which were used for selecting significant shared features (combined p<0.05).

In addition to overall models, we conducted exploratory descriptive analyses stratified by clinical diagnosis background (ASD lines: ASD_HNRNPU_, ASD_CASK_; non-ASD lines: CTRL_Male_, CTRL_Female_) to identify potential trends in exposure responses. Given the small number of lines per stratified group, these analyses were not powered for formal interaction testing.

### Pathway enrichment analysis

Detected metabolites were tested for enrichment in the Small Molecule Pathway Database (SMPDB)^39^ using the quantitative enrichment analysis (QEA) approach to metabolite set enrichment analysis (MSEA) from the enrichment analysis module of MetaboAnalyst 5.0.^38^ Any missing values were imputed using the in-built K-Nearest Neighbour (KNN) algorithm for similar samples. The metabolite labels of 172 out of 189 metabolites were successfully mapped to PubChem (https://pubchem.ncbi.nlm.nih.gov) and annotated with PubChem CIDs. The remainder (17) were excluded from the analysis. The concentration data was normalised using data-scaling (auto-scaling, as done in the feature selection). The cut-off threshold for entries from metabolite sets to be included from the metabolite set library was set to 2.

QEA uses a generalised linear model to estimate a *Q-statistic* for each metabolite set, which describes the correlation between compound concentration profiles, X, and experimental outcomes, Y. The *Q statistic* for a metabolite set is the average of the *Q statistics* for each metabolite in the set. The cut-off threshold for entries from metabolite sets to be included was set to 2. Based on the statistical testing, enrichment dot-plots and pathway networks were generated. In the pathway networks, 2 or more nodes were only connected by an edge when the number of shared metabolites was >25% of the combined metabolites contributing to each node.

### Lipid exposure and cPLA2 inhibition

Solutions of the selected LPCs (LPC 16:0, 855675P, Sigma-Aldrich; LPC 18:0, 845875P, Sigma-Aldrich; and LPC 18:1, 855775P, Sigma-Aldrich) were prepared in methanol (stock: 0.01 M; 322415, Sigma-Aldrich) and used immediately by warming up at 37°C or stored at - 20°C for future use. Working solutions were prepared based on the determined concentration of the LPCs at day 5 of SH exposure (LPC 16:0 – 2.0 µM, LPC 18:0 – 1.8 µM and LPC 18:1 – 0.8 µM), from the mass spectrometry based-metabolomics (Fig. 3A). Vehicle controls were also prepared, as required. The cells were exposed to a 2X mixture of the LPCs at the initiation of undirected differentiation and every media change (alternative feeding days). LPC synthesis was inhibited by tandemly exposing the differentiating cells with 10.0 µM AACOCF_3_ (arachidonyl trifluoromethyl ketone; ab120350, Abcam). AACOCF_3_ is a phospholipase-modulating compound with reported PLA2-inhibitory activity, and was used to perturb LPC-related lipid pathways.^40–42^ Cellular ROS and cellular ATP assays were performed at day 5 and 28 of exposure, as described in the previous sections. For these assays, two independent experiments were performed for each differentiation timepoint, with a suitable number of technical replicates as visualised in the plots. All readings were reported as percentage of control. Two-way ANOVA followed by Tukey’s post-hoc correction for multiple comparisons, was used to test for significance (adj. p<0.05) of the reported findings in R (v4.1.2).^30^

### Population-based mother-child cohort

The Barwon Infant Study (BIS) consists of 1074 mother-child pairs (including 10 sets of twins) recruited from the Barwon region (Victoria, Australia) recruited antenatally between July 2010 and July 2013. Cohort details, including population characteristics, eligibility and exclusion criteria, and data collection methods, have been described elsewhere.^43^ The BIS cohort is broadly representative of the Australian population, with a slight over-representation of families from English-speaking backgrounds.^43^ Detailed cohort characteristics relevant to the present analyses are provided in Supplementary Table S6A.

Information on antidepressant use, including the type used, was collected in a medication questionnaire during the 28-week antenatal review (1st and 2nd trimester). In addition, the Edinburgh Postnatal Depression Scale (EPDS)^44^ was administered during the 28-week antenatal review. This is a widely used and validated measure of depression that assesses mood over a one-week period. Further details are described earlier.^45^

Lipidomic measures were obtained from cord blood serum sample (obtained from the umbilical cord blood at birth, then placed into serum clotting tubes and analysed with ultra-high-performance-tandem mass spectrometry (UHPLC-MS/MS). The time from blood collection to processing and storage, as well as the total duration of storage at -80°C, was recorded and considered in the subsequent analysis. Maternal blood contamination in cord blood samples was assessed using DNA methylation profiling based on previously established methods.^46,47^

The details of the UHPLC-MS/MS analysis are described elsewhere.^48^ Briefly, 776 lipid features were quantified, including LPC 16:0, LPC 18:0, and LPC 18:1, with the fatty acyl moiety at either the sn1 or sn2 position. These were chosen *a priori* based on the results of the *in-vitro* experiments. Lipids were extracted using a 1:1 butanol solution with 10 mM ammonium formate, which included a mixture of internal standards. The analysis was conducted using UHPLC-MS/MS on an Agilent 6490 QQQ mass spectrometer coupled with an Agilent 1290 series UHPLC system. Separation was achieved using two ZORBAX Eclipse Plus C18 columns (21 × 100 mm, 18 mm, Agilent) at a temperature of 45°C. Mass spectrometry was performed in both positive and negative ion modes with dynamic scheduled multiple reaction monitoring. Lipid species concentrations were determined by comparing them to the corresponding internal standards. Further details, data processing and quantification have been described previously.^46,48^

ADHD and autism-related symptoms were assessed using the Child Behaviour Checklist for ages 1.5–5 (CBCL) at 2 years and the Strengths and Difficulties Questionnaire P4-10 (SDQ) at 4 years, methods described elsewhere.^46^ Specifically, the DSM-5-oriented subscales for attention-deficit/hyperactivity problems (CBCL-ADHP) and autism spectrum problems (CBCL-ASP) from the CBCL were utilised, along with the SDQ subscales for hyperactivity (SDQ-hyper), peer problems (SDQ-peer), and prosocial behaviour (SDQ-prosocial). The questionnaires were completed by the child’s parent or guardian.

Multivariate regression models were applied to assess the association between prenatal SSRI use, antidepressant use, and EPDS score and LPC levels in cord blood. Multivariate regression models, with LPCs split into categories based on quartiles, to allow for nonlinear associations with dose-response and neurodevelopmental outcomes, were also assessed. Covariates included child sex, gestational age, time sample spent on the bench and in the freezer before analysis, contamination of the child’s blood with maternal blood, and child’s birth weight. LPC-neurodevelopmental outcomes were additionally adjusted at the child’s age the CBCL or SDQ was completed. Nominal statistical significance was considered at p<0.05 and trends at p<0.10.

### Statistical analyses

All statistical analyses were performed in R (v4.1.2).^30^ The statistical models and tests used for the analyses, along with any inclusion/exclusion criteria are described in the methodology relevant to the experimental technique, in the sections above. All plots, unless otherwise stated, were created using the *ggplot2* package (v3.3.5)^37^ or the *pheatmap* package (v1.0.12, https://CRAN.R-project.org/package=pheatmap) in R (v4.1.2).^30^

## RESULTS

### Increased comparative cytotoxicity is observed with sertraline

We studied the *in-vitro* effects of four SSRIs, namely fluoxetine hydrochloride (FH), citalopram hydrobromide (CH), sertraline hydrochloride (SH) and paroxetine hydrochloride (PH) using four human iPSC derived NES cell lines, of which two were from individuals with NDDs and known pathogenic genetic variants.^25–28^ NES cells underwent undirected differentiation for up to 28 days, with and without SSRI exposure.

First, we tested the optimal concentrations for SH, CH, and PH using the CTRL_Male_ cell line, similar to our prior work with FH.^23^ Using an exposure range between 0.0–30.0 µM, cell viability was assessed using the MTS assay at 24- and 120-hours post-differentiation. The highest concentration with no significant reduction in viability (Tukey post hoc p>0.05) or observable changes in cellular morphology under phase-contrast microscopy at day 5 was selected. Based on these criteria, the optimal concentrations were 3.0 µM for FH, CH, and PH and 1.5 µM for SH (Supplementary Figure S1A). SH demonstrated increased cytotoxicity at 120 hours post-differentiation compared to the other SSRIs, for which its optimal concentration was adjusted.

### Sertraline and paroxetine decrease cellular ROS and ATP levels

Total ROS levels were detected using the DCFDA (2’,7’-dichlorodihydrofluorescein diacetate) assay at day 5 and 28 of differentiation in the four cell lines. At day 5 and day 28, a significant (Nemenyi post hoc p=0.005) reduction in ROS levels was detected for SH exposure (Fig. 1A). For PH, significantly reduced total ROS levels were detected at day 28 (Nemenyi post hoc p=0.007) (Fig. 1A).

**Fig. 1.**
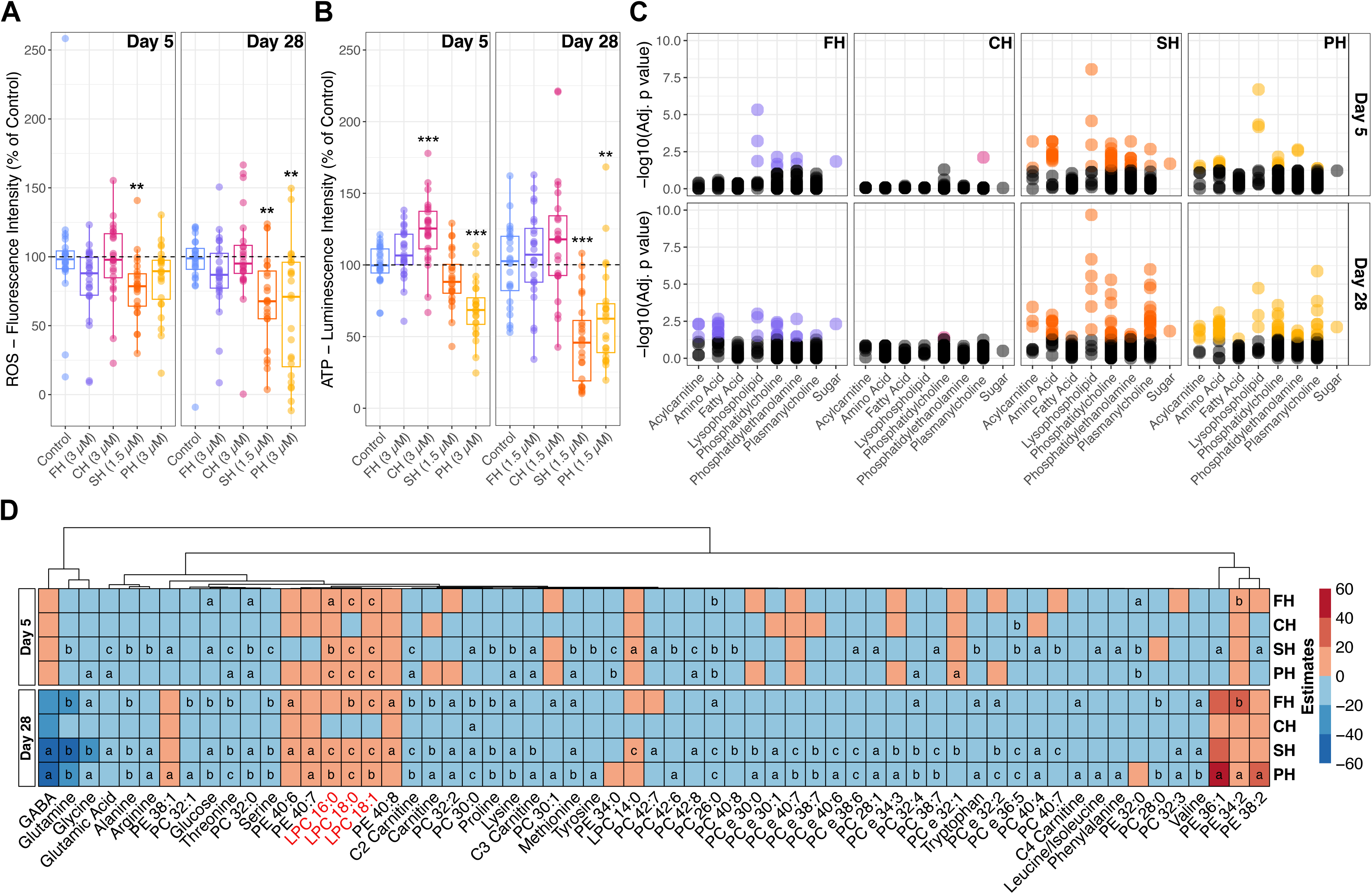
*In-vitro* assays and differential metabolomics following selective serotonin reuptake inhibitor (SSRI) exposure in differentiating neuroepithelial stem cells. **A.** *In-vitro* assay determined total ROS levels at day 5 (n=24 per condition) and day 28 (n=24 per condition) of fluoxetine (FH), citalopram (CH), sertraline (SH) and paroxetine (PH) exposure (Kruskal-Wallis Rank Sum, Nemenyi post hoc p: *<0.05, **<0.01, ***<0.001). B. *In-vitro* assay determined total ATP levels at day 5 (n=24 per condition; two-way ANOVA, Tukey’s post hoc adjusted p: *<0.05, **<0.01, ***<0.001) and day 28 (n=24 per condition; Dunn (1964) Kruskal-Wallis, Holm post hoc p: *<0.05, **<0.01, ***<0.001) of FH, CH, SH and PH exposure. **C.** Distribution of significant metabolites across metabolite classes at day 5 and day 28 (mixed linear model, FDR adjusted p). **D.** Heatmap of estimate values from the applied mixed linear model for metabolites and SSRIs, with a significant overlap across at least two SSRIs (FDR adjusted p: a<0.05, b<0.01, c<0.001).

To correct for the Crabtree Effect, where cells in culture prefer glycolysis over oxidative phosphorylation,^31^ the cells were assayed in a cell medium in which glucose was replaced with galactose to promote ATP generation by oxidative phosphorylation (Supplementary Figure S1B). CH significantly increased (Tukey post hoc p=0.0004) total ATP levels, while PH caused a decrease (Tukey post hoc p<0.0001) at day 5 (Fig. 1B). At day 28, no difference was observed with CH compared to the control; however, both SH (Holm adj. p<0.0001) and PH (Holm adj. p=0.005) decreased total ATP levels (Fig. 1B).

### Early metabolic changes are induced by sertraline

Mass spectrometry-based untargeted metabolomics revealed over 2500 metabolite signals, including adducts, in our cell lysates. Of these, 189 unique metabolites met the signal threshold and were quantified at both day 5 and day 28 post neural differentiation and SSRI exposure. At day 5, cells were exposed to FH, CH, and PH at 3.0 µM and SH at 1.5 µM, and the exposure was validated in the metabolomics data (Supplementary Figure S1C). PCA revealed overlapping metabolic profiles across the SSRI treatments, with PC1 and PC2 accounting for 43.5% of the total variability (32.6% and 10.9%, respectively) (Supplementary Figure S2A). Herein, the SSRI mediated effects are not driven by large-scale metabolomic shifts but specific metabolite changes which can be visualised using volcano plots (Supplementary Figure S3A).

Using a mixed linear model accounting for the genetic background and the exposure followed with FDR correction, we report that SH modulated the largest number of metabolites (52 metabolites), followed by PH (19 metabolites), FH (9 metabolites), and CH (1 metabolite) at day 5 (Supplementary Table S2A). For SH, the metabolites spanned seven classes: acylcarnitines, amino acids, lysophospholipids, phosphatidylcholines, phosphatidylethanolamines, plasmanylcholines and sugars (Fig. 1C). Amino acids such as glutamic acid (SH, adj. p=0.001; PH, adj. p=0.019), methionine (SH, adj. p=0.004; PH, adj. p=0.027), threonine (SH, adj. p=0.001; PH, adj. p=0.014), and lysine (SH, adj. p=0.004; PH, adj. p=0.049) were reduced by both SH and PH (Supplementary Table S2A and Fig. 1D). Feature selection analysis of day 5 data highlighted ten significant metabolites across SSRI treatments (Supplementary Table S2D). The lysophosphatidylcholines – LPC 16:0, LPC 14:0, and LPC 18:0 emerged as the top three features, each with a combined p-value of <0.0001.

### Paroxetine and sertraline share metabolic profiles following prolonged exposure

For the day 28 metabolomics analysis, all four cell lines were treated with SSRIs at a standardized concentration of 1.5 µM, based on the SH treatment level. These exposures were confirmed in the samples (Supplementary Figure S1C). Again, PCA did not show clear clustering by treatments (Supplementary Figure S2B). PC1 and PC2 explained 38.1% of the variance (22.8% and 15.3%, respectively). Furthermore, specific metabolite changes can be visualised using volcano plots (Supplementary Figure S3B).

The mixed linear model analysis detected the highest number of metabolite changes after PH treatment (64 metabolites), followed by SH (61 metabolites), FH (28 metabolites), and CH (1 metabolite) (Supplementary Table S3A). Most metabolites affected by SH, FH, and CH were also modulated by PH (Fig. 1C). For instance, PC 30:0 was significantly reduced by all SSRIs (FH, adj. p=0.004; CH, adj. p=0.040; SH, adj. p=0.018; PH, adj. p=0.0001). Amino acids such as glutamine (FH, adj. p=0.007; SH, adj. p=0.003; PH, adj. p=0.005), proline (FH, adj. p=0.004; SH, adj. p=0.003; PH, adj. p=0.004), and threonine (FH, adj. p=0.002; SH, adj. p=0.001; PH, adj. p=0.001) were decreased across these treatments (Fig. 1D). GABA was specifically reduced by SH (adj. p=0.023) and PH (adj. p=0.035) (Supplementary Table S3A and Fig. 1D).

LPCs (18:0, 18:1, 16:0) were consistently elevated: LPC 18:0 (FH, adj. p=0.002; SH, adj. p<0.0001; PH, adj. p<0.0001), LPC 18:1 (FH, adj. p=0.001; SH, adj. p<0.0001; PH, adj. p=0.001), and LPC 16:0 (SH, adj. p<0.0001; PH, adj. p=0.008) (Fig. 1D). Feature selection using metabolite data from day 28 identified 24 significant metabolites (Supplementary Table S3D). The top three features were LPC 18:0 (combined p<0.0001), LPC 18:1 (combined p=0.001), and LPC 16:0 (combined p=0.002).

### Exploratory analysis of diagnosis-related SSRI exposure effects

To complement the primary analyses of SSRI exposure effects across all four cell lines, we examined responses stratified by the clinical background of the cell donors to identify exploratory diagnosis-related patterns in cellular readouts and metabolomics. This analysis revealed only modest diverging patterns. In terms of total ROS levels, a significant decrease was observed at day 28 in the non-ASD lines following SH (Nemenyi post hoc p=0.022) and PH (Nemenyi post hoc p=0.002) exposures (Supplementary Figure S4A). At day 5, both non-ASD and ASD lines showed increased total ATP with CH (non-ASD, Tukey post hoc p=0.004; ASD, Tukey post hoc p=0.040), while PH decreased total ATP in both groups, more prominently in the ASD lines (non-ASD, Tukey post hoc p=0.017; ASD, Tukey post hoc p=0.0002) (Supplementary Figure S4B). At day 28, sertraline caused a significant total ATP decline in both groups, with a larger reduction in the non-ASD lines (non-ASD, Holm adj. p=0.007; ASD, Holm adj. p=0.019) (Supplementary Figure S4B).

In the metabolomic signature analysis, across timepoints, the ASD lines exhibited fewer changes at Day 5 (SH: 18, CH: 4, FH: 2, PH: 1; Supplementary Table S2B and Figure S4D), when compared with the non-ASD lines (SH: 24, PH: 10, FH: 6, CH: 0; Supplementary Table S2C and Figure S4C). However, at Day 28 this pattern reversed, with the ASD lines showing more extensive modulation (SH: 55, PH: 29, FH: 18, CH: 0; Supplementary Table S3B and Figure S4F) relative to the non-ASD lines (SH: 17, PH: 14, FH: 5, CH: 0; Supplementary Table S3C and Figure S4E). Some diverging patterns also emerged, for instance, fatty acids FA 10:0 and FA 20:1 were increased, while PC 26:0 was decreased only in the ASD lines, with CH and SH at Day 5 (Supplementary Table S2B and Figure S4D). At Day 28, GABA decreased only with SH in both the ASD and non-ASD lines (Supplementary Table S3B and S3C), and the short-chain acyl-carnitine, C3 carnitine decreased with SH and PH (Supplementary Table S3B and S3C, Figure S4E and S4F). Overall, these exploratory comparisons largely support shared lipid effects, with limited diagnosis-specific features.

### Phospholipid biosynthesis and amino acid metabolism are key pathways modulated by SSRI exposure

Using metabolite-set enrichment analysis (MSEA) and quantitative enrichment analysis (QEA), significant biochemical pathways affected by SSRI exposures were identified. At day 5, exposure to FH (Fig. 2A, Supplementary Table S4A) resulted in the enrichment of ten pathways, including phospholipid biosynthesis (adj. p=0.0001), galactose metabolism (adj. p=0.0001), and sphingolipid metabolism (adj. p=0.001). SH exposure significantly enriched 48 pathways (Supplementary Table S4C), notably phospholipid biosynthesis (adj. p<0.0001), purine metabolism (adj. p=0.001), and tryptophan metabolism (adj. p=0.001) (Fig. 2A and Supplementary Figure S2C). Similarly, PH exposure enriched 50 pathways (Supplementary Table S4D), including phospholipid biosynthesis (adj. p<0.0001), purine metabolism (adj. p=0.014), and tryptophan metabolism (adj. p=0.027) (Fig. 2A and Supplementary Figure S2D). In contrast, CH exposure did not result in significant pathway enrichment (Supplementary Table S4B).

**Fig. 2.**
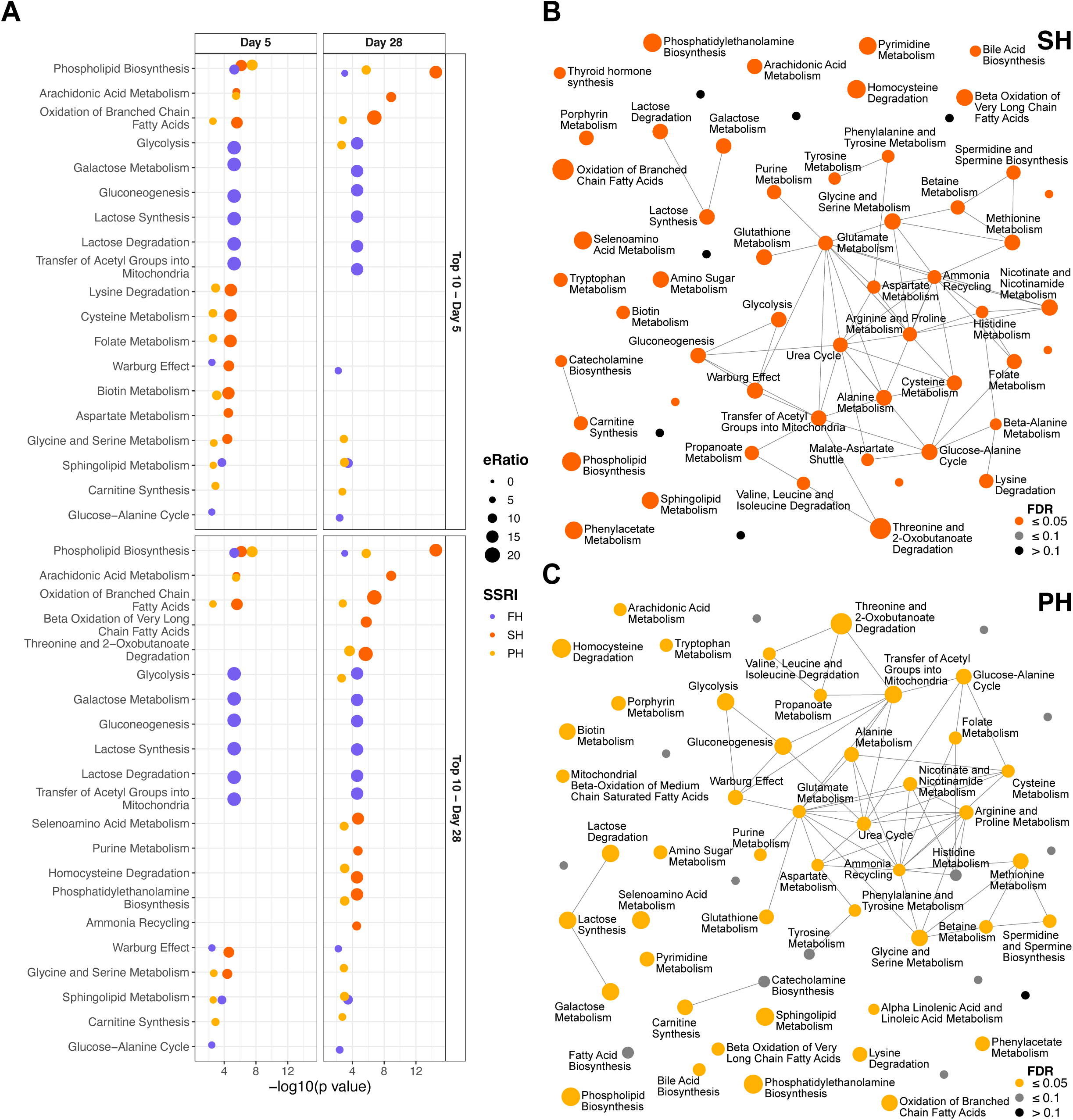
Enrichment of detected metabolites in biochemical pathways. **A.** Enrichment dot-plot of top 10 significantly enriched metabolic pathways (adjusted p ≤0.05) following fluoxetine (FH), sertraline (SH) and paroxetine (PH) exposure at day 5 and day 28. **B.** Interaction network of top 50 enriched metabolic pathways following SH exposure at day 28. **C.** Interaction network of top 50 enriched metabolic pathways following PH exposure at day 28.

At day 28, similar results were observed. FH exposure enriched ten pathways (Supplementary Table S5A), such as galactose metabolism (adj. p=0.0003), sphingolipid metabolism (adj. p=0.003), and phospholipid biosynthesis (adj. p=0.007) (Fig. 2A). SH exposure led to the enrichment of 54 pathways (Supplementary Table S5C), including phospholipid biosynthesis (adj. p<0.0001), purine metabolism (adj. p=0.0002), and phenylalanine and tyrosine metabolism (adj. p=0.003) (Figs. 2A and 2B). PH exposure enriched 46 pathways (Supplementary Table S5D), including phospholipid biosynthesis (adj. p=0.0001), purine metabolism (adj. p=0.018), and phenylalanine and tyrosine metabolism (adj. p=0.041) (Figs. 2A and 2C). CH exposure continued to show no significant pathway enrichment (Supplementary Table S5B).

These findings emphasize the consistent enrichment of key biochemical pathways, particularly phospholipid biosynthesis and amino acid metabolism, in response to FH, SH, and PH exposures. This highlights the central role of these pathways in SSRI-induced metabolic effects.

### Selected LPCs associate with *in-vitro* and *in-vivo* SSRI exposures

As LPC 18:0, LPC 18:1, and LPC 16:0 were the most consistent findings within the cellular exposure metabolic screening at both time points (Figs. 1D and 3A), we continued to investigate their effects both *in-vitro* and *in-vivo*. First, we hypothesized that LPCs could contribute to changes in ROS and ATP levels that were observed after the SH and PH exposures (Figs. 1A and 1B). To test this, we inhibited LPC production using the phospholipase A2 inhibitor arachidonyl trifluoromethyl ketone (AACOCF_3_, 10 µM) for 24 hours, as well as treated the cells with a combination of the LPCs (18:0, 18:1, 16:0) and SH. The 24-hour inhibition of the LPC production attenuated the reduction of ROS both at day 5 and day 28 in the SH-treated cells; however, there was no significant difference between the two conditions (Fig. 3B). For ATP production, the LPC production inhibition only slightly attenuated the effect of SH on day 5 and seemed to enhance the reduction on day 28 (Fig. 3C). Interestingly, the combination of LPCs increased the ROS and ATP levels at both time points (Supplementary Figures S2E and S2F). Our findings indicate that the three LPCs and inhibition of their synthesis were associated with changes in ROS and ATP levels, though inconsistently, suggesting more complex regulatory patterns.

**Fig. 3:**
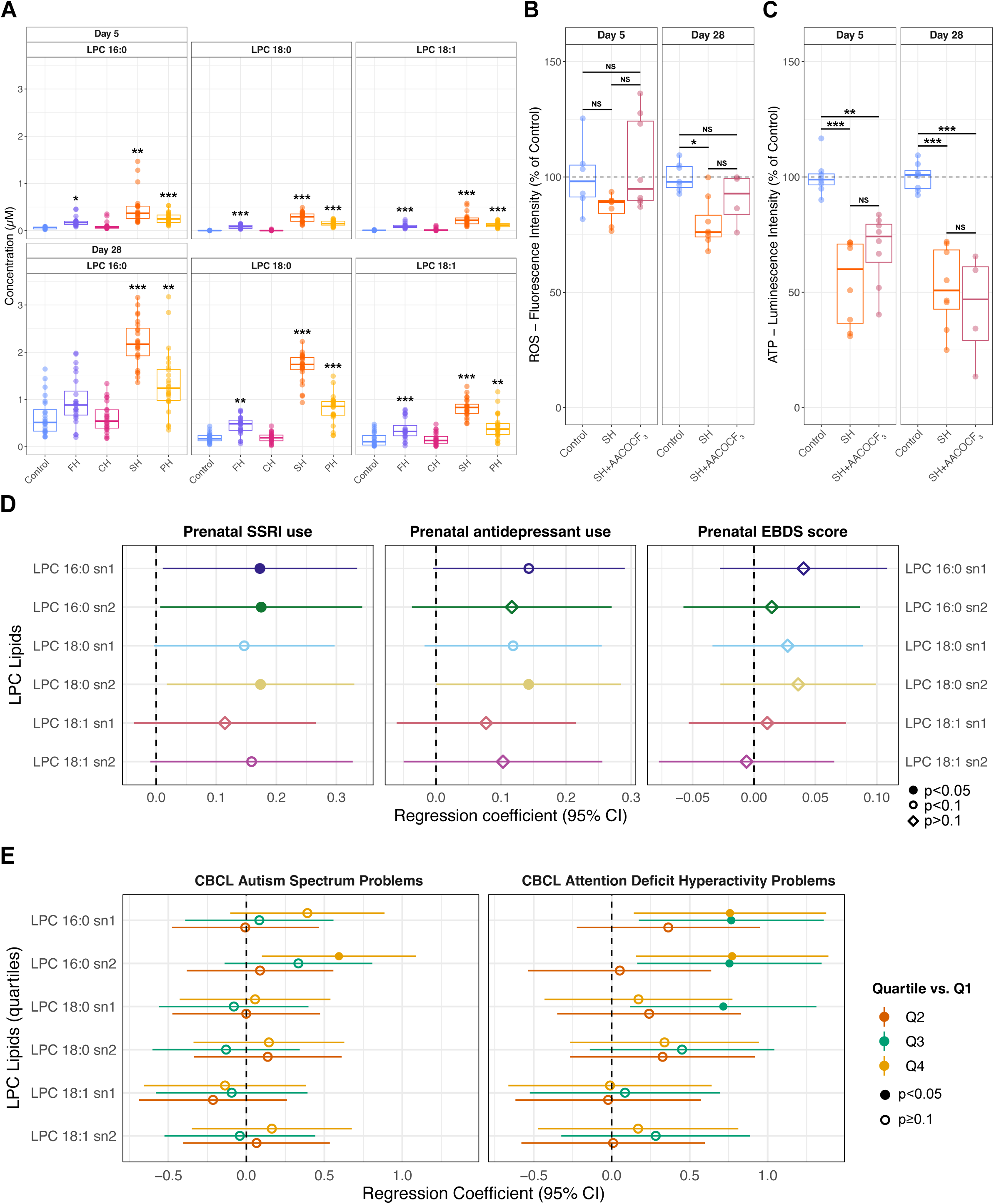
Association of selected lysophosphatidylcholines. (**LPCs) with selective serotonin reuptake inhibitor (SSRI) exposures and neurodevelopmental disorder outcome measures. A.** Detected concentration of selected LPCs following fluoxetine (FH), citalopram (CH), sertraline (SH) and paroxetine (PH) exposure at day 5 (n=24 per condition) and 28 (n=24 per condition) (mixed linear model, FDR adjusted p: *<0.05, **<0.01, ***<0.001). B. *In-vitro* assay determined total ROS levels at day 5 (Control, n=6; SH, n=8; SH+AACOCF_3_, n=8) and day 28 (Control, n=7; SH, n=8; SH+AACOCF_3_, n=4) of LPC inhibition (two-way ANOVA, Tukey’s post hoc adjusted p: *<0.05, **<0.01, ***<0.001; NS: non-significant). C. *In-vitro* assay determined total ATP levels at day 5 (n=8 per condition) and day 28 (n=8 per condition, except SH+AACOCF_3_, n=4) of LPC inhibition (two-way ANOVA, Tukey’s post hoc adjusted p *<0.05, **<0.01, ***<0.001; NS: non-significant). **D.** Adjusted estimated mean change in doubling of LPC concentration (with 95% confidence interval, CI), per exposure. **E.** Adjusted estimated mean change in CBCL (Child Behaviour Checklist) Autism Spectrum Problems and CBCL Attention Deficit Hyperactivity Problems (with 95% CI) by quartiles of LPC concentrations (Q1, reference group).

Next, we used metabolic and neurodevelopmental outcome data from the BIS cohort,^46^ comprising 1,074 mother-child pairs, to analyse the LPC effects in a population-based cohort. First, we investigated the associations between maternal SSRI use (n=40) and any antidepressant use (n=48) within the first and second trimesters on the levels of the selected LPCs (Fig. 3D, Supplementary Table S6B) in the infant cord blood. Our findings indicate that prenatal SSRI use was associated with elevated levels of cord blood LPC 16:0 sn1 (β=0.173, 95% CI=0.011–0.334; p=0.037), and LPC 16:0 sn2 (β=0.175, 95% CI=0.007–0.343; p=0.042), and LPC 18:0 sn2 (β=0.174, 95% CI=0.018–0.330; p= 0.029). Additionally, we observed trends (p<0.1) suggesting higher levels of LPC 18:0 sn1 and LPC 18:1 sn2 with prenatal SSRI use. When analysing the effects of any antidepressant use, which added eight additional mothers on other antidepressants, these associations were attenuated (Fig. 3D, Supplementary Table S6B). There were no significant correlations between maternal prenatal EPDS scores and any of the LPCs studied (Fig. 3D, Supplementary Table S6B). Furthermore, no significant associations were found between prenatal SSRI use and maternal LPC levels at 28 weeks of gestation or with the infant’s LPC levels at 6 months, 12 months, and 4 years.

We previously published significant positive associations between LPCs and CBCL autism and ADHD symptom scales, although not all remained significant after FDR correction.^46^ Here, we further examined the relationship between the three consistently upregulated LPCs and the outcomes by categorizing each LPC measure into quartiles. This approach allowed us to explore potential non-linear associations with CBCL-autism and CBCL-ADHD scores and assess dose-response relationships. Significant associations were found for LPC 16:0 sn2 with CBCL-autism scores (overall p=0.01) and for both LPC 16:0 sn1 and LPC 16:0 sn2 with CBCL-ADHD scores (overall p=0.007 and 0.002, respectively) (Fig. 3E, Supplementary Table S6C).

When investigating outcomes older than two years of age, we did not observe significant associations between any of the LPCs and autism/ADHD-related measures from the strength and difficulties questionnaire (SDQ) at age 4 for hyperactivity difficulties, peer problem difficulties, prosocial behaviour strengths, or parent-reported autistic traits (Supplementary Table S6D).

## DISCUSSION

We have investigated the effects of four commonly prescribed SSRIs on human iPSC-derived neuroepithelial stem cells undergoing undirected differentiation for 5 and 28 days towards neurons using four genetically distinct human iPS cell lines. Exposure to SH and PH, of which the latter is not clinically recommended during pregnancy,^49^ notably decreased total ROS and ATP levels, compatible with altered oxidative phosphorylation, though direct mitochondrial function was not assessed. The metabolomic analysis further identified multiple significantly modulated metabolites predominantly driven by SH and PH, enriched in energy metabolism and amino acid pathways. Importantly, we identified elevated lysophosphatidylcholines (LPC 16:0, LPC 18:0, and LPC 18:1) as robust molecular signatures consistent across cellular models and clinical samples, underscoring their potential clinical significance. The clinical validation using the population-based BIS cohort confirmed the LPC 16:0 and LPC 18:0 as potential candidate biomarkers correlating with prenatal SSRI exposure. Our findings suggest LPCs may be associated with SSRI exposure and neurodevelopmental traits, but further validation in larger cohorts is required.

There is a lack of clarity regarding the ability of SSRIs to alter ROS levels and their mechanism of action. The association between SSRIs and ROS has been reported predominantly in epidemiological and *in-vivo* studies of depression.^50–53^ In clinical depression, the antioxidant systems are known to be inherently suppressed^19,54^ suggesting that the antioxidant, ROS-reducing properties of SSRIs apply only under oxidative stress.^19,55^ We hypothesise that reduced ROS availability during key neurodevelopmental stages could influence ROS-mediated signalling pathways that are important for neurite growth and synaptogenesis.^21^ We also highlighted differences in *in-vitro* ATP levels, which we use as a surrogate marker for normal mitochondrial function. Both SH and PH have been previously shown to induce mitochondrial damage with depleted ATP levels, but in astrocytes.^56^ Another *in-vitro* study on human placental cells also suggests that SH alters ROS and ATP levels.^57^ While our results align with some observations reported in the literature, the precise, underlying mechanisms by which SSRIs affect ROS and ATP levels remain largely unclear.

In addition to ROS and ATP levels, we detected several pathways mediated by FH, SH, and PH exposure related to energy metabolism. The SSRIs, specifically escitalopram (enantiomer of CH) and FH, have been shown to localize in cellular phospholipid membranes, cytoplasm, and endoplasmic reticulum, influencing various aspects of cellular physiology.^24^ Interestingly, in human immortalised cell lines of brain origin, FH was demonstrated to activate the AMPK-ACC-CPT1 pathway,^58^ which is known to play an important role in energy-related metabolic homeostasis through the regulation of mitochondrial ROS.^59^ FH has also been shown to enhance energy metabolism in the hippocampus of an *in-vivo* model of major depressive disorder.^60^ A comprehensive multi-omics study has also supported the link between FH and increased energy metabolism across multiple rat brain regions.^61^ FH exposure in mice has been shown to induce the gradual accumulation of sphingomyelin,^62^ a pathway we highlighted in our earlier study^23^ and this study. While there are fewer studies with CH, SH, and PH in this domain, CH has been known to cause *in-vitro* phospholipidosis,^63^ and PH has been reported to cause energy metabolism changes in animal models.^64–66^

We also reported enrichments in pathways related to the metabolism of different amino acids crucial for normal brain function. In an animal model, PH demonstrated an impact on metabolites involved in amino acid metabolism.^65^ This is significant as we have previously shown differences in urinary metabolites derived from amino acids in individuals diagnosed with autism.^67^ Others have also reported an association of tryptophan, purine, phenylalanine, and tyrosine metabolism with autism.^68–72^ Intriguingly, CH did not seem to have any major effects on cellular metabolism in our study despite a recent report that prenatal exposure is linked to higher rates of ADHD diagnosis.^73^ We acknowledge the requirement for more work with higher doses of CH to further investigate its effects *in-vitro*.

In addition to the enrichment of metabolic pathways, our main finding was the consistent increase of the three LPCs (LPC 16:0, LPC 18:0, and LPC 18:1) following SSRI exposure in neural cells and in cord blood. While these observations suggest altered phospholipid metabolism, the underlying mechanisms remain unclear. LPC 18:0 has been demonstrated to be an agonist for the peroxisome proliferator-activated receptor-gamma (PPAR-γ) pathway in a mouse model.^74^ In the brain, the PPAR-γ pathway has been connected with motivation and associative learning.^75^ Treatment of rodent models of autism with an agonist of the PPAR-γ pathway was shown to improve several phenotypic features.^76,77^

We also reported a decrease in the levels of several phosphotidylcholines, which has been previously associated with autism^78^ and can be attributed to an increased activity of cytosolic phospholipase A2 (cPLA2).^78,79^ The enzyme cPLA2 hydrolyses phospholipids to LPCs and other bioactive lipids,^80^ potentially contributing to the observed increase in LPC levels that we have reported. Since, cPLA2 activity and PPAR-γ signalling were not measured in this study, further research is required to connect these downstream effects with SSRI exposure that could potentially lead to premature neural differentiation^81^ during early development.

We need to acknowledge that elevated LPC levels in the cord blood are not the only metabolites associated with autism and ADHD traits as there were several other metabolites associated with these outcomes in the BIS cohort.^46^ However, pending further investigation, this could be more specific for *in-utero* SSRI exposure and thus related to a broad number of outcomes also reported in the offspring. For instance, higher levels of LPC 18:0 in maternal serum have been indicated as a biomarker for congenital heart defects (CHDs) in infants.^82^ Furthermore, LPC 16:0 and LPC 18:1 levels in cord blood were positively associated with birth weight,^83–85^ a known liability factor for different cardiometabolic conditions.^86^ This, in turn, could be connected with PH, as its early *in-utero* exposure has been correlated with an increased liability of cardiac malformations in infants.^49,87^ There is a link between CHDs and adverse neurodevelopmental outcomes,^88,89^ which warrants further investigation of the *in-utero* SSRI exposure effects through the LPCs. Additionally, *in-utero* SSRI exposure was found to increase anxiety and depression-related symptoms in adolescents, along with changes in the activation of fear-related brain circuitry.^90^ Nevertheless, it is also essential to highlight that despite potentially adverse indicators of SSRI-mediated effects *in-utero*, a recent large-scale study demonstrated a reduction in depression-related mental health outcomes following postnatal SSRI treatment, with subsequent benefits observed on child externalising behaviours during early childhood as well.^91^

Although our study has many strengths, we must also recognise its limitations. Despite employing four different cell lines, the small cell line panel size limits our ability to distinguish biological heterogeneity from stochastic variability, and we lacked sufficient power to make strong comparisons regarding sex or genetic background in relation to exposure effects, indicating a need for further research. Nevertheless, we have accounted for putative genetic differences in our findings by also including cell lines derived from individuals with NDDs and reported the exploratory analyses of putatively different exposure outcomes based on clinical diagnoses status of the iPSC-donors. Additionally, our *in-vitro* system does not model maternal pharmacokinetics, placental transfer, foetal metabolism, or maternal mental health, and the SSRI concentrations, therefore, represent cellular exposure paradigms rather than exact foetal brain or serum levels.

While the BIS cohort provides a robust clinical validation for our *in-vitro model*, its observational design is susceptible to confounding factors, including genetic influences. Additionally, the relatively small number of infants exposed to SSRIs (n=40) limits the generalisability of our findings. Furthermore, BIS is broadly representative of the Australian population, though families from English-speaking backgrounds are slightly over-represented,^43^ which should be considered when extrapolating the findings. Furthermore, the antidepressant exposure in BIS was ascertained by maternal self-report via a medication questionnaire completed at the 28-week antenatal review. This approach may introduce self-reporting and recall inaccuracies. Although prospective reporting reduces differential recall bias, misclassification is possible and is likely non-differential, which would bias exposure–outcome associations toward the null. Dosage, adherence, and medication use outside the reporting window were not captured and could contribute to exposure heterogeneity. We also need to acknowledge that our study did not cover the issue of SSRI withdrawal, which has been associated with neuroinflammation in a murine model and a potential elevation of ROS levels.^92^ The metabolomic and neurological impact of withdrawal in early life, either at or prior to weaning, also requires evaluation. Overall, larger studies are required to determine whether LPC 16:0 and LPC 18:1 levels in cord blood statistically mediate an association between maternal SSRI use and autism-related outcomes in infants. In terms of policy, the advantages and disadvantages of SSRI use in pregnancy requires continual revaluation as new findings, such as those described here, occur.

In conclusion, this comprehensive translational investigation provides novel insights into the metabolomic impact of prenatal SSRIs on human neuronal differentiation and neurodevelopment. We identify LPC 16:0 and LPC 18:0 as promising candidate metabolites of altered lipid metabolism, potentially mediating the relationship between prenatal SSRI exposure and increased likelihood of autism and ADHD traits. Future studies should clarify the molecular mechanisms underlying these metabolomic alterations and assess their clinical utility in liability prediction and targeted interventional strategies.

## CONTRIBUTIONS

Conceptualization: A.A., P.V., A-L.P. and K.T.; Methodology: A.A., P.V., A-L.P. and K.T.; Software development: A.A., K.V. and K.T.; Validation: A.A., K.V., C.M., M.-O.D. F.M., J.H., X.Y. and M.O.; Formal Analysis: A.A., K.V., C.M., M.-O.D., F.M. and J.H.; Investigation: A.A., K.V., C.M., M.-O.D., F.M., J.H., X.Y. and M.O.; Resources: P.V., A-L.P., I.L. and K.T.; Data Curation: A.A., K.V., C.M. and F.M.; Writing – Original Draft: A.A., M.-O.D, J.H. and K.T.; Writing – Review & Editing: A.A. and K.T.; Visualization: A.A., K.V. and K.T.; Supervision: A-L.P., I.L., and K.T.; Project Administration: A.A. and K.T.; Funding Acquisition: A.A. and K.T. All authors have read and approved the final version of the manuscript. A.A. and K.T. have accessed and verified the underlying data of the manuscript. The members of the Barwon Infant Study Investigator Group were Mimi L.K. Tang, Lawrence Gray, Sarath Ranganathan, Peter Sly, Martin O’Hely, Richard Saffery, David Burgner and Len Harrison.

## ACKNOWLEDGMENTS

The authors would like to thank the donors and their parents for providing valuable material for this study as a part of the RATSS (the Roots for Autism and ADHD Twin Study in Sweden) cohort. The authors acknowledge the RATSS team, especially Prof Sven Bölte, Dr Karl Lundin Remnelius, Dr Johan Isaksson and Dr Janina Neufeld for their support. The authors also acknowledge the BIS cohort participants and assistance from Lada Holland for data cleaning and verification. We thank Prof Peter J. Meikle, Dr Satvika Burugupalli and the Metabolomics laboratory at the Baker Institute for the conduct and advice on lipidomic lab measures for BIS.

## ETHICS

The Swedish Ethical Review Authority (2016/1452-31 and 2012/208-31/3) gave ethical approval for the research conducted at Karolinska Institutet. The Barwon Infant Study was approved by the Barwon Health Human Research and Ethics Committee (HREC 10/24). Written informed consent was obtained from all participants or their caregivers, based on their age.

## DATA AVAILABILITY

The data and code required for differential metabolomics analysis are available on GitHub (https://github.com/Tammimies-Lab/SSRI-Metabolomics). The raw data from the mass spectrometry-based metabolomics will be available on MetaboLights upon publication of the preprint or available upon request from the corresponding author. The data from the Barwon Infant Study (BIS) are available under restricted access to protect participant confidentiality. Researchers may request access by contacting the BIS Steering Committee via Anne-Louise Ponsonby at the Florey Institute of Neuroscience and Mental Health. Requests are considered based on scientific and ethical considerations, and approved applications require a collaborative research agreement. Deidentified data can be shared in either Stata or CSV file formats. Further details about the cohort, data access process, and project overview are available at: www.barwoninfantstudy.org.au.

## SUPPLEMENTARY INFORMATION

Supplementary information can be found as an appendix to the preprint.

## Supplementary figures

**Supplementary Figure S1:**
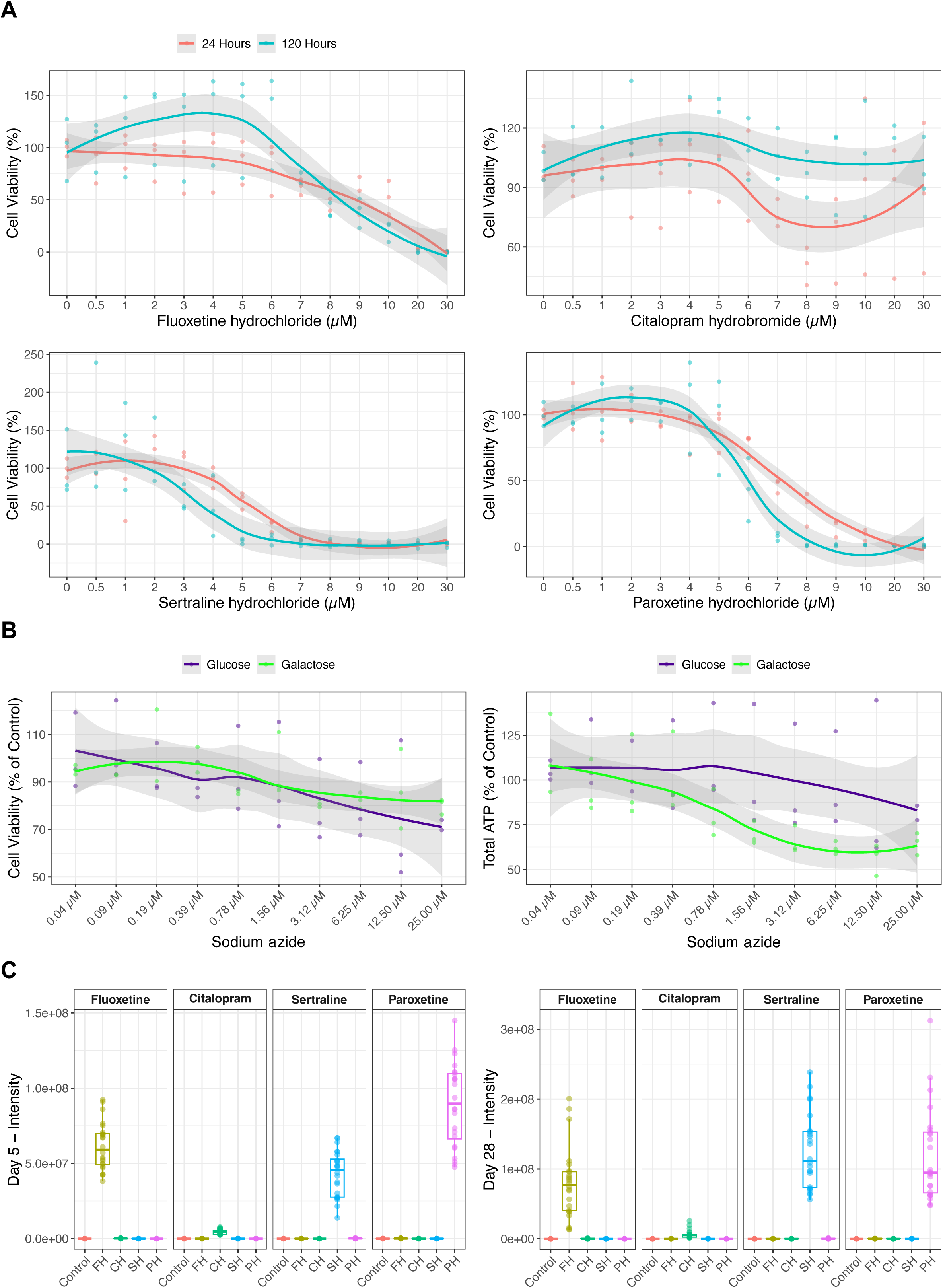
Cellular assays for method optimisations. **A.** Cell viability at 24 hours (n=3 per condition) and 120 hours (day 5, n=3 per condition) of fluoxetine (FH; from Arora et al., 2023, Scientific Reports), citalopram (CH), sertraline (SH) and paroxetine (PH) exposure using the MTS assay. **B.** Optimisation of the shift from glycolysis to oxidative phosphorylation using glucose free, galactose supplemented medium and testing with sodium azide exposure using a mitotoxicity assay (cell membrane permeability, n=3 per condition and total ATP, n=3 per condition). **C.** Detection of FH, CH, SH, and PH in the mass spectrometry readout following *in-vitro* exposure at day 5 (n=24 per condition) and 28 (n=24 per condition).

**Supplementary Figure S2:**
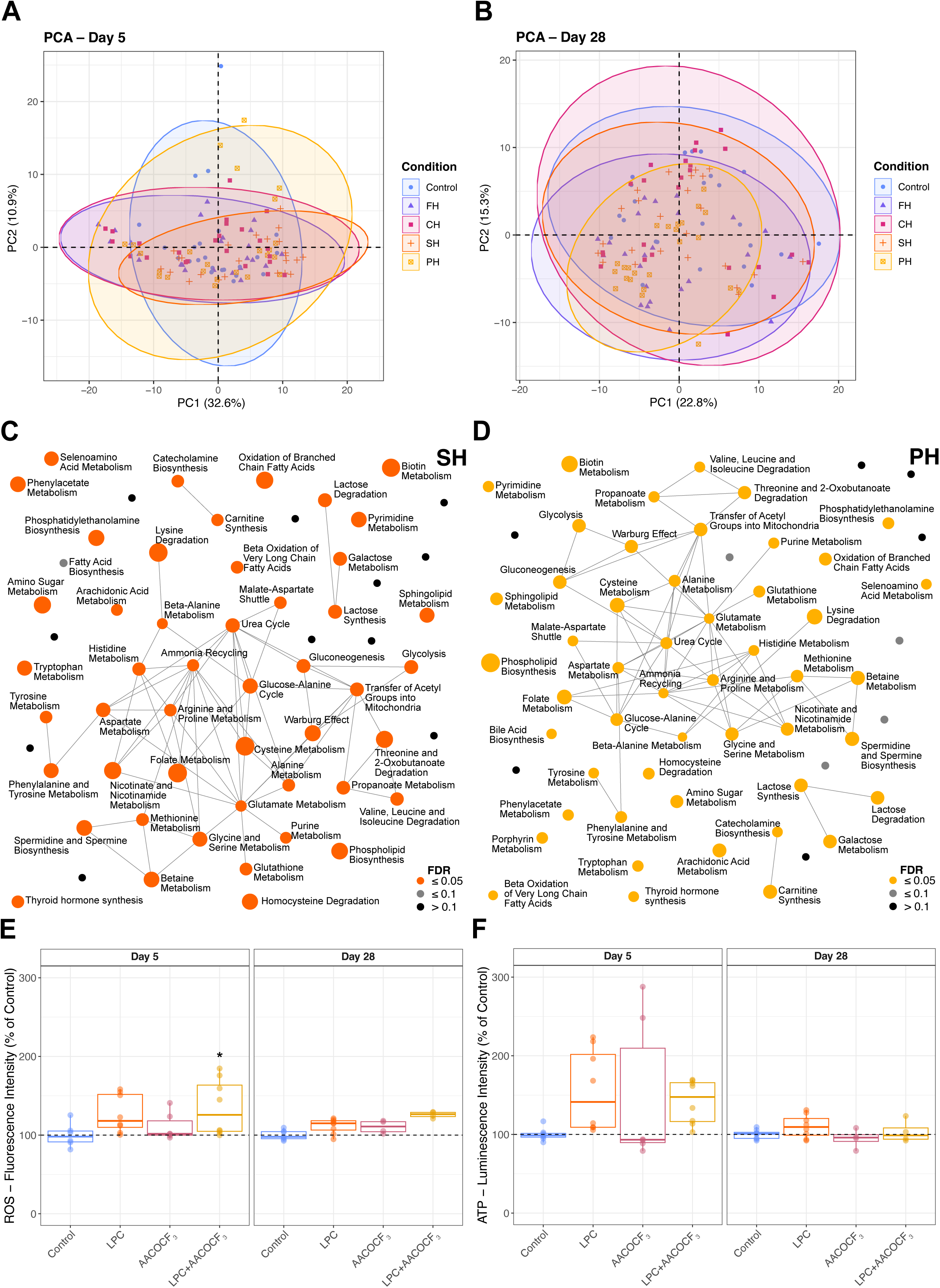
Analysis of selective serotonin reuptake inhibitor (SSRI) exposure effects. **A.** Principal component analysis (PCA) scatter plot of detected metabolites following fluoxetine (FH), citalopram (CH), sertraline (SH) and paroxetine (PH) exposure at day 5 (n=24 per condition). **B.** PCA scatter plot of detected metabolites following FH, CH, SH and PH exposure at day 28 (n=24 per condition). **C.** Interaction network of top 50 enriched metabolic pathways following SH exposure at day 5. **D.** Interaction network of top 50 enriched metabolic pathways following PH exposure at day 5. **E.** *In-vitro* assay determined total ROS levels at day 5 (Control, n=6; lysophosphatidylcholines, LPC, n=8; AACOCF_3_, n= 6; LPC+AACOCF_3_, n=8) and day 28 (Control, n=7; LPC, n=8; AACOCF_3_, n= 4; LPC+AACOCF_3_, n=4) of LPC inhibition (two-way ANOVA, Tukey’s post hoc adjusted p: *<0.05, **<0.01, ***<0.001). **F.** *In-vitro* assay determined total ATP levels at day 5 (Control, n=8; LPC, n=8; AACOCF_3_, n= 6; LPC+AACOCF_3_, n=8) and day 28 (Control, n=8; LPC, n=8; AACOCF_3_, n= 4; LPC+AACOCF_3_, n=4) of LPC inhibition (two-way ANOVA, Tukey’s post hoc adjusted p *<0.05, **<0.01, ***<0.001).

**Supplementary Figure S3:**
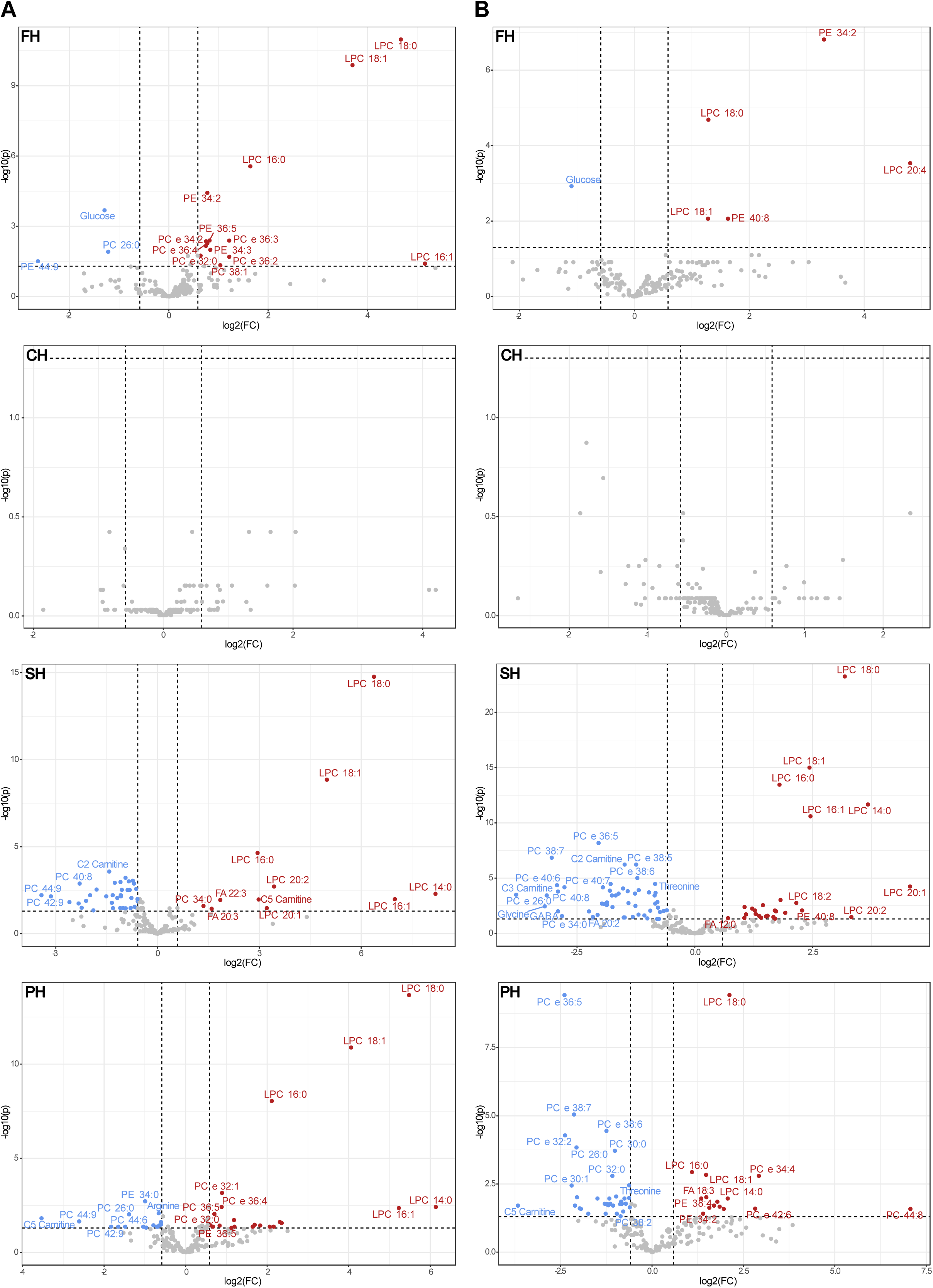
Univariate analysis of selective serotonin reuptake inhibitor (SSRI) exposure effects. **A.** Volcano plots for differentially detected metabolites following SSRI exposure at day 5. **B.** Volcano plots for differentially detected metabolites following SSRI exposure at day 28.

**Supplementary Figure S4:**
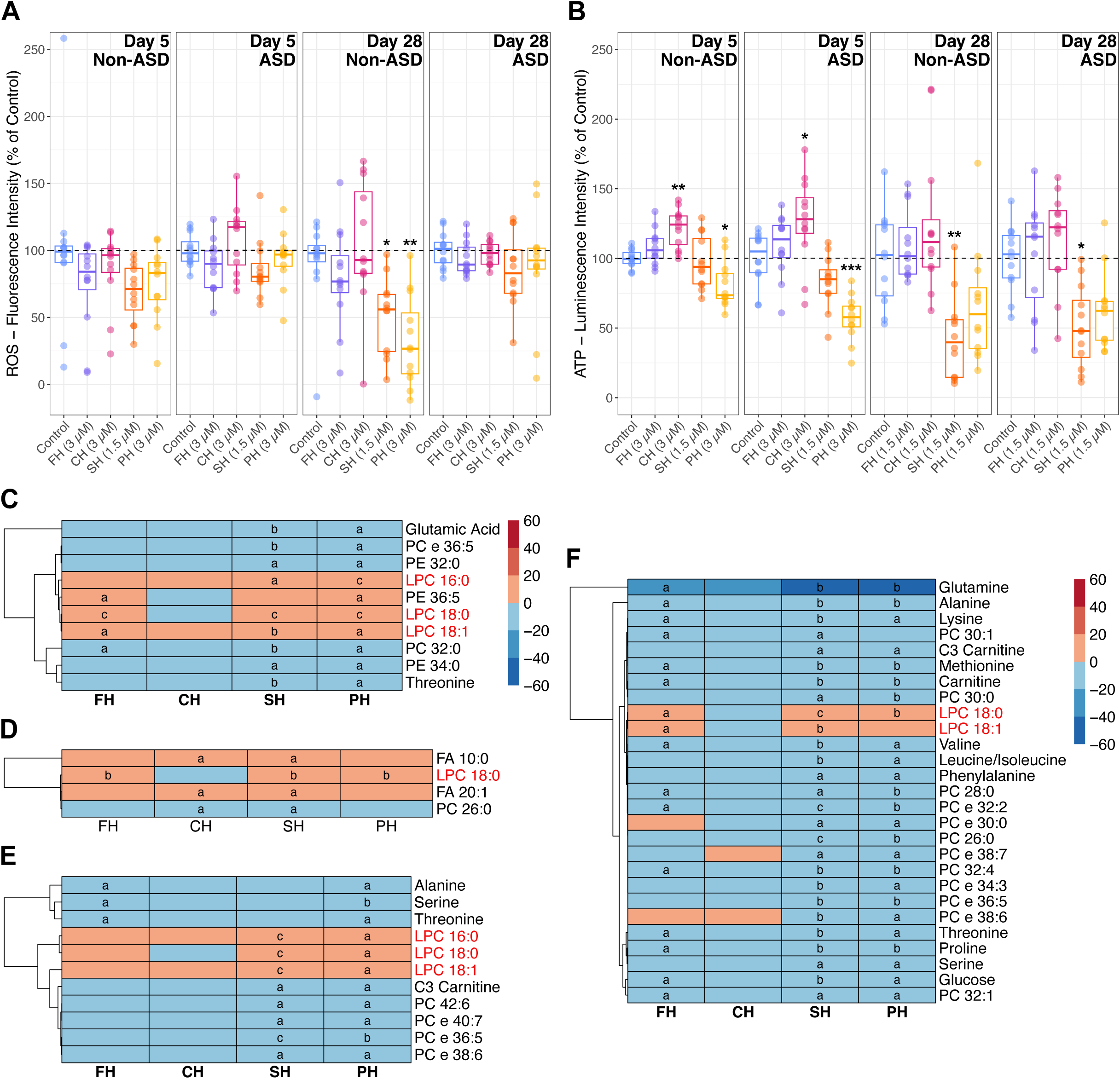
Selective serotonin reuptake inhibitor (SSRI) effects as per clinical diagnostic group. **A.** *In-vitro* assay determined total ROS levels at day 5 (n=12 per condition) and day 28 (n=12 per condition) of fluoxetine (FH), citalopram (CH), sertraline (SH) and paroxetine (PH) exposure in the non-ASD and ASD lines (Kruskal-Wallis Rank Sum, Nemenyi post hoc p: *<0.05, **<0.01, ***<0.001). **B.** *In-vitro* assay determined total ATP levels at day 5 (n=12 per condition; two-way ANOVA, Tukey’s post hoc adjusted p: *<0.05, **<0.01, ***<0.001) and day 28 (n=12 per condition; Dunn (1964) Kruskal-Wallis, Holm post hoc p: *<0.05, **<0.01, ***<0.001) of FH, CH, SH and PH exposure in the non-ASD and ASD lines. **C.** Heatmap of estimate values from the applied mixed linear model for metabolites and SSRIs, with a significant overlap across at least two SSRIs in the non-ASD lines at Day 5 (FDR adjusted p: a<0.05, b<0.01, c<0.001). **D.** Heatmap of estimate values from the applied mixed linear model for metabolites and SSRIs, with a significant overlap across at least two SSRIs in the ASD lines at Day 5 (FDR adjusted p: a<0.05, b<0.01, c<0.001). **E.** Heatmap of estimate values from the applied mixed linear model for metabolites and SSRIs, with a significant overlap across at least two SSRIs in the non-ASD lines at Day 28 (FDR adjusted p: a<0.05, b<0.01, c<0.001). **F.** Heatmap of estimate values from the applied mixed linear model for metabolites and SSRIs, with a significant overlap across at least two SSRIs in the ASD lines at Day 28 (FDR adjusted p: a<0.05, b<0.01, c<0.001).

## SUPPLEMENTARY TABLE

**Supplementary Table S1.** Characteristics of induced pluripotent stem cell lines included in the study.

| Published Name | Name in Study | RRID | Cell Type | Source Organism | Sex | Clinical Background | QC & Source Ref. |
| --- | --- | --- | --- | --- | --- | --- | --- |
| AF22 | CTRL<sub>Female</sub> | CVCL_A5CW | iPSC | *Homo sapiens* | Female | Neurotypical Control | Falk et al., 2012 (PLOS One) |
| CTRL-9-II | CTRL<sub>Male</sub> | CVCL_JL74 | iPSC | *Homo sapiens* | Male | Neurotypical Control | Uhlin et al., 2017 (Stem Cell Research) |
| HNRNPU<sub>del/+</sub> | ASD<sub>HNRNPU</sub> | N/A | iPSC | *Homo sapiens* | Male | ASD with *HNRNPU* heterozygous deletion | Mastropasqua et al., 2023 (Biology Open) |
| ASD<sub>CASK_SS</sub> | ASD<sub>CASK</sub> | N/A | iPSC | *Homo sapiens* | Male | ASD with *CASK* splice-site variant | Becker et al., 2020 (Translational Psychiatry) |

RRID: Research Resource Identifier, N/A: Not Available, iPSC: Induced Pluripotent Stem Cell, ASD: Autism Spectrum Disorder, *HNRNPU*: Heterogeneous nuclear ribonucleoprotein U, *CASK*: Calcium/Calmodulin Dependent Serine Protein Kinase, Ref.: Reference.

